# Educational interventions to combat burnout: Are General Practitioners interested? A Qualitative Study

**DOI:** 10.1101/2024.03.27.24304944

**Authors:** Darius Williams

## Abstract

**Background:** Rising burnout in General Practitioners has been noted as a major issue worldwide that is contributing to many leaving the profession earlier than anticipated or reducing their hours of clinical work. Educational interventions as part of continuing professional development (CPD) have been shown to be effective in reducing burnout and improving overall wellbeing amongst GPs. There is no published evidence available describing attitudes of GPs toward such CPD-based burnout interventions. The objective of this study is to assess GPs’ perspectives and opinions towards educational CPD-based interventions aimed at improving burnout in the GP population.

**Methods:** A qualitative research approach using grounded theory methods was used. Participants were interviewed 1 to 1 via video call. Video files of the interviews were recorded and auto-transcription software used to generate a text file which was checked for accuracy of transcription. Transcripts underwent grounded thematic analysis with emergent themes synthesised and combined to develop a targeted analysis concordant with the objectives of the study. The study received ethical approval from the Swansea University Medicine, Health and Life Science Ethics Approval Board.

**Results:** 5 participants were interviewed – all were GPs listed on the GMC GP register and currently engaged in the Wales GP appraisal process. Two participants had prior experience of CPD resources focused on burnout. Participants universally noted positive sentiment towards an educational-based CPD intervention focused at reducing burnout and noted their preferences in how such an intervention might be designed. Several important perceived barriers were highlighted that would need to be considered in the design of any future interventions.

**Discussion:** Attitudes of participants suggest an educational-based CPD intervention would be well received, and further research is needed to assess the efficacy and cost-effectiveness of such an intervention in this population.

## Introduction

Burnout has been defined as “a psychological syndrome characterized by emotional exhaustion, depersonalization and a reduced sense of professional efficacy that can appear in caregivers”. (Maslach, 1979; Maslach & Leiter, 2016) It is an increasingly recognised phenomenon in multiple occupations and population groups such doctors (Schaufeli et al., 2009), social workers (Martínez-López et al., 2021), teachers (Pressley, 2021) and carers of young children. (Griffith, 2022) This has led to burnout being recognised as a specific occupational disorder in the 11^th^ version of the WHO’s International Classification of Diseases (ICD-11). (World Health Organisation, 2019)

Rising burnout in GPs has been noted as a major growing issue that is contributing to many leaving the profession earlier than anticipated or reducing their hours of clinical work. (British Medical Association, 2015; Dale et al., 2015, 2016; Iacobucci, 2021) Hall et al demonstrated, in a survey of 232 UK based GPs, 93.8% were classed as likely to be suffering from a minor psychiatric disorder with 72.7% respondents reporting severe exhaustion. (Hall et al., 2019) Orton et al similarly demonstrated high levels of emotional exhaustion (46%) and depersonalisation (42%) and low levels of personal accomplishment (34%) in a survey-based study of 564 UK-based GPs. (Orton et al., 2011) This compares with two global meta-analyses of burnout in GPs - one performed by Shen et al which demonstrated that 37% and 28% of 7595 GPs suffer from high levels of emotional exhaustion and high depersonalization, respectively. (Shen et al., 2022) Another by Karuna et al demonstrated rates of high emotional exhaustion (32%) and high depersonalisation (31%) in the 22,177 GPs sampled. (Karuna et al., 2022) Burnout has also been shown to result in poorer outcomes and quality of care for patients. (Hall et al., 2019; Iliffe & Manthorpe, 2019; Salyers et al., 2017) This represents a major problem in the context of an ageing population and an already stretched healthcare system within the United Kingdom.

Several groups have sought to assess the utility of CPD/educational interventions in improving rates of burnout in General Practitioners. (Asuero et al., 2014; Barcons et al., 2019; Fortney et al., 2013; Gardiner et al., 2004, 2013; Krasner et al., 2009; Margalit et al., 2005; Montero-Marin et al., 2018; Schroeder et al., 2018; van Wietmarschen et al., 2018) These studies used varying educational interventions including cognitive behavioural stress management training (Gardiner et al., 2004, 2013), mindfulness based educational interventions varying from a few weeks (Asuero et al., 2014; Montero-Marin et al., 2018; Schroeder et al., 2018; van Wietmarschen et al., 2018) to up to a year (Krasner et al., 2009), biopsychosocial focused training via didactic/interactive sessions (Margalit et al., 2005) and a multimodal training programme with an Integrated Brief Systemic Therapy combined with a mental health support programme. (Barcons et al., 2019) Collectively these studies clearly demonstrate that educational interventions as part of CPD can help to reduce burnout and improve overall wellbeing amongst GPs.

Whilst there is evidence to support the use of educational CPD interventions to help prevent burnout in the GP population, there is a dearth of evidence that GPs in Wales have had any exposure to such interventions. It is also unclear whether these interventions are something that GPs feel they want or need. Given the mandatory requirement for CPD in the UK, it would potentially represent a useful opportunity to provide interventions to improve GP wellbeing and, consequently, patient outcomes.

The objectives of this study are twofold:

1. To discover if GPs in South Wales have had any exposure to/awareness of programmes providing CPD/educational interventions to help reduce burnout or improve resilience whilst exploring any relevant experiences of these, or other non-CPD based interventions of which participants may be aware.
2. To assess whether GPs would find a CPD/educational intervention focused at improving burnout helpful, enquiring how GPs would want such a scheme to run, exploring possible formats and perceived barriers.

## Method

A qualitative research approach using grounded theory methods was used to examine GPs experiences and preferences with regard to CPD, alongside their experiences of CPD-based interventions to prevent burnout or improve wellbeing/resilience in the workplace. A qualitative approach was chosen in preference to a quantitative method, such as a survey-based assessment, as it would give a more detailed understanding of the experiences of GPs and allow for exploration of reasoning in greater depth. The Consolidated Criteria for Reporting Qualitative Research (COREQ) checklist was followed. (Tong et al., 2007)

### Ethical Approval

The study received ethical approval from the Swansea University Medicine, Health and Life Science Ethics Approval Board.

### Recruitment

All GPs listed on the GMC GP register and engaged in the Wales GP appraisal process were eligible for recruitment in the study. A purposive sampling strategy was used for recruitment. It is acknowledged that the varying routines and day-to-demands upon GPs in different career roles (eg partners versus portfolio GPs) are likely to mean that perspectives on and methods of accessing CPD were likely to be very different. In light of the smaller sample sizes used in a qualitative study, it is likely that a randomly chosen sample may miss out on these varied perspectives. As such, the author decided, where possible, to select examples of each role in order to capture these varying perspectives.

Recruitment emails were sent to potential participants with a patient information sheet and consent form attached. All study documents received prior ethical approval from the Swansea University Medicine, Health and Life Science Ethics Approval Board. Upon recruitment, participants were informed of the anonymity procedures used in the study and all questioned answered prior to signing of the consent form. Following completion of the interview, a second check was undertaken prior to data anonymisation to ensure that participants were in agreement with the collected data being included in the study.

### Data Collection

Interviews were performed by DW between August and September 2023. Either in-person or remote video call options offered, depending on the preference of the participant. Interviews lasted typically 30 minutes. A topic list was created by the authors to give a degree of uniformity to the interview process. However, an iterative approach was taken with the interview questions with additions and refinements made based on the topics and themes that developed during the interviews. The aims of the study and possible goals were explained to participants during the informed consent process. No non-participants were present during the interview process and no field notes were created during the interview process.

### Reflexivity

DW is a male General Practitioner (MBBCh MRCGP) with a special interest in GP wellbeing, working in the South Wales area in an Academic GP role. This is spent part-time in clinical practice and part-time in research/education roles. The participants are current or ex-colleagues of DW. This may introduce a source of selection bias and the author acknowledges that the nature of an interview where the participant has a prior relationship with the interviewer may potentially yield different results when compared with an unknown interviewer. It was felt that a prior relationship would lead to a more authentic and honest conversation, with the participants more easily able to explore sensitive issues surrounding burnout.

### Data Analysis

Video files of the interviews were recorded and auto-transcription software via Microsoft Office/Teams was used to generate a text transcription of the interview. (Microsoft, 2023b, 2023a) This text file was then reviewed and compared by the author to the recorded conversation to ensure accuracy of transcription. Transcripts were not returned to participants after transcription. Following this, transcribed text files were imported to Nvivo qualitative research software. (Lumivero, 2018) They then underwent grounded thematic analysis with emergent themes synthesised and combined to develop a targeted analysis concordant with the objectives of the study. (Braun & Clarke, 2006, 2012; Cooper et al., 2012; Kennedy & Lingard, 2006; Tavakol et al., 2006; Watling & Lingard, 2012) All analysis of transcripts was performed by DW. After analysis of 5 transcripts, data saturation was felt to have been reached. Participants were not contacted to provide feedback on the findings of the study.

## Results

### Participant Demographics

Participants were General Practitioners, registered on the GMC GP register, currently practicing in Wales and engaged in the All Wales GP Appraisal Process. All participants who received recruitment emails agreed to participate. All participants fully completed the interview process, signing the consent form prior to any data being collected and additionally agreeing that their anonymised data could be included, after the interview had been completed.

Five participants were interviewed in total. All interviews were undertaken via video call using MS Teams. (Microsoft, 2023a) No repeat interviews were performed. Age of participants ranged from 32 years to 54 years (median = 41 years, mean = 40.8 years). Three participants described their gender as female, and two as male. One participant identified their job role as senior GP partner, one as salaried GP, one as portfolio GP, one as academic GP and one as locum GP. Years practicing as a GP ranged from three years to 25 years (median = 7 years, mean = 11 years). Participants described their main location of clinical work as Cardiff and Vale University Health Board, Cwm Taf Morgannwg University Health Board or Swansea Bay University Health Board.

Three themes were extracted from the data via grounded thematic analysis. These are shown in Table 1.

**Table 1:** Themes.

|  |
| --- |
| 1) General Sentiments |
| 2) Design Preferences |
| 3) Specific Barriers & Concerns |

### Combatting burnout – Experiences of CPD and non-CPD based approaches

Two participants had previously undertaken an educational activity that focused on recognition or management of burnout/work-related stress. These were generic, online or in-person taught modules. These participants found these educational interventions of partial benefit, providing basic conceptual training in recognition of burnout symptoms which was helpful, but they highlighted the need for a more bespoke, flexible approach to such interventions. The other participants had neither undertaken nor heard of any such interventions in the community or with colleagues:

> *Yeah, it was an online module. Again, back to online stuff, and I think it was with the BMJ and I think it was something I thought “uh, that’s probably is quite relevant to you” and I think it was on, umm, stress I think. You know, I was going through a difficult time myself in terms of just being in practice I was unhappy with. So I think I I looked at this to, sort of, again maybe…just as a going back to this showing and demonstrating, so that like when I went to appraisal “it’s been quite a difficult year and do you know what, I’ve done this, I’ve done this module” (Participant 3)*
>
> *It was quite generic really, you know and I think it needs to vary based on situations. It needs to be individualized, almost. (Participant 3)*
>
> *I think I did. I was asked to attend a leadership course pre-COVID, I think it was 2017 when clusters were fairly new cluster leads were, you know, asked to go on leadership courses. I attended the very first course on leadership. And there was, I think there was eight modules to that course over a six-month period in XXX and run by HEIW. And that was good, all about Myers-Briggs and looking at, you know, your own personality and the personalities around the cluster tables et cetera. And I think one module included burnout. So that’s kind of CPD type of thing on burnout and how to recognise it and out to and how to mitigate against it, you know. (Participant 5)*
>
> *I can understand the need to have a generic “Here’s a recognition of burnout*.*” You know, as an organization, at an organizational level, you sort of sort of, for example, Cardiff University or the NHS, you know you need to be able to provide people with the tools to identify it (Participant 3)*

Several participants were aware of other interventions that GPs had undertaken to try to improve stress levels and stave off burnout. These included a regular lunchtime yoga session, a regular positive feedback practice email letter and use of podcasts discussing burnout issues:

> *I know that a friend of mine did do some CPD on burnout for her practice. Soon after qualifying she became a partner and during COVID she set up, actually, I think, I don’t know where she got the resources from, possibly from the RCGP, but she basically set up kind of a lunchtime kind of meditation yoga session that she used to take, and, for her practice was like on different sites essentially. So she would set it up and then they would like whoever could join could join into that. And I think they found that helpful as a way to kind of stay connected through COVID with people, but also have a bit of time where they could just like, sit and relax. (Participant 1)*
>
> *The other thing actually that I heard that, um, one of my colleagues did, which was kind of related, I suppose, to burnout was actually they used to send round a like a happy newsletter every month that would, like, collate all of the positive comments that people would make, like, patients would make. And because a lot of the time, they’re not written down because people often say nice things verbally but would potentially put complaints in writing and not always put nice things in writing, so morale was quite low in the surgery. What they did, they set it up and everyone would send anything nice or any, you know, any compliments, essentially, of staff to her. And then every month it would get sent out. So everyone could kind of see it in writing, the nice things that people said. And I thought that was a really nice idea, actually, to kind of boost morale. (Participant 1)*
>
> *No, I vaguely remember, you know, using the Facebook groups a lot. There’s a couple of podcasts that I think that focus on it that a couple of the GPs on the groups have directed us to but I’ve never accessed them myself. (Participant 5)*

Although not specifically enquired about in the interview, several participants shared some of their own experiences with burnout and discussed their own routines they had developed to help manage work-based stresses:

> *I think everyone’s different. Everyone’s got their own level of resilience that you know, for me, I run now every day or do something every day or climb something and that is…that’s what I do and I have to do that to feel…just to get that headspace. And that’s my resilience. (Participant 2)*
>
> *I’ve had days where I’ve felt burnout, if that makes sense. It’s never been, you know, and for me, what it feels like on those days is that I’m just angry that people are there. Feeling just like, “I don’t know why you’re here” and just or being angry that someone hasn’t…I don’t know…when they say “oh, I did think about calling the ambulance about this” you know, why didn’t you? You know, like just having that sort of inner feeling like you just actually…you don’t care (Participant 2)*
>
> *I’m certainly working in an area which would make…and we’ve got failed practices around us that have gone to the wall. So that tells you something about the area. So I would agree with that. In terms of mental exhaustion, fatigue and things, I think I’m definitely close to burnout (Participant 4)*

### CPD-based interventions for burnout

#### 1. General Sentiments

When asked to reflect on their feelings surrounding usage of an educational intervention to improve burnout and enhance resilience, participants felt this was a novel and potentially beneficial idea. None of the participants expressed any negative attitudes towards the concept of an education-based intervention:

> *I would be happy to engage in them. I think they’re really valuable and it*..*oh yeah, I don’t really, I think the only real barrier would be finding the time I suppose to do them and a lot of them, like meditation, are things you’d need to be doing regularly. So you’d have to get into a routine of doing it and I think that’s probably the biggest barrier is just getting yourself into that routine. But yeah, I’d be happy to do it and I think it is valuable. (Participant 1)*
>
> *I do think it’d be a good idea. I think lots of people would benefit from it. (Participant 2)*
>
> *I think, as long it’s adequately resourced, I can see that being beneficial. (Participant 4)*
>
> *Yeah, yeah, why not? You know, I think I would be more open to that, if that came into the e-mail inbox or was like a, you know, this is a course that’s offered, yeah, yeah, rather than just more general broad, sort of, burnout and ways to become more resilient. Because there’s also that question of like, well, you’re treating the problem, you’re treating the problem and not the cause or however that saying goes. So you know what everyone’s going to be is, like, “well, I’m burnt out because the system’s rubbish and, you know, I wouldn’t be if I could just do what I need for my patients”. But that’s the life, isn’t it? It’s not going to change anytime soon. So you might as well have some techniques to tackle it. (Participant 5)*

#### 2. Design Preferences

Participants were asked to consider how they felt an educational intervention to focus on burnout would be best designed. The majority of participants felt that a group-based intervention would be the optimum design:

> *I think group work or, you know, group based CPD may be helpful. I think people tend to…it’s like the Facebook groups, you know, like resilient GP or GP survival. People like to vent as a group. And I think, not that this would be the arena for it necessarily, but I think that can be really helpful and therapeutic. I think, it’s what it’s like with any sort of therapy isn’t it, not that I’m suggesting this is therapy! But yeah, you know, if we’re speaking to people in a similar situation and we’re having a similar time, being able to vent, discuss experiences, offload. If that was a facet of it, I think that would be helpful. So yeah, maybe something more face to face, group based (Participant 5)*
>
> *I can see it working better in groups. It could be an economy of scale as well. And to do that, I think you need, as you say you need a sort of central bank of doctors that can go into a practice, because that practice has released their GP for that day to go on this course with another 30 odd other GPs (Participant 4)*
>
> *it needs to be a group or an individual based small groups, individual…needs to be face to face or online, there needs to be that contact with people doesn’t it really, to be able to deliver an educational intervention like that around this type of topic (Participant 3)*
>
> *I think maybe like when we said with psychiatry having those access to those type of groups where if you need to go and talk to people like, yeah, if you knew in your area in your cluster or I don’t know, there was a group that ran once a month or something, you could go to it and actually talk with other GPs and there was someone there who was, I don’t know, specially, to actually give you some advice about how to manage stress, how to manage burnout, some sort of, yeah, like not like a drop in, but yeah, somewhere where you could just go. And there were other GPs because I think actually having other people saying, yeah, I’m, I’m going through that too…it’s really powerful to do things in groups, because you feel like you’re probably the only one going through it. So I think more of a group based thing would be useful and maybe some online. You know, if you that you could tap in and out of, yeah (Participant 2)*

It was also felt that, although remote courses would potentially overcome geographical barriers and have improved scalability, an intervention dealing with complex psychological issues was felt to be most appropriate when given face to face:

> *I think if it was online it probably just wouldn’t be done because, like I said, I think it would just be at the bottom of the pile for people to do and so I think some sort of like face to face thing can make it interactive and also maybe kind of be able to talk to other people, I guess about the risk of burning out and people in a similar position to you (Participant 1)*
>
> *I would think it would probably be better done in more of a face to face way and probably in fairly small chunks, cause I think people wouldn’t want to give up a huge amount of time just because that in itself would kind of be, I suppose, stressful for them to have to give up a huge amount of time. (Participant 1)*

Opinion surrounding an e-module based design was varied. Some participants saw the value in a separate stand-alone module which gave basic degree of education as to what burnout is and along with basic strategies to manage symptoms with some additional signposting to further resources. Others felt an e-module to be an inappropriate design for this topic:

> *I think it’s stages, isn’t it? You have that standalone module and then then the next stage might be that you have a group session (Participant 3)*
>
> *Not an e-module. You know, despite me having said like, that’s typically what I do for appraisal, I can’t…I don’t know whether I would sort of seek that out or access that, in that scenario (Participant 5)*

One participant felt that, whilst there was benefit to this intervention in GPs, a similar intervention should be aimed at medical colleagues at earlier stages in their career, such as specialist training level or even at an undergraduate level:

> *if something like that happens, then you feel like the rug’s been pulled from under you and so I think if there was some type of teaching preparing you maybe, maybe as a GP trainee or even before to recognize what it what it is, recognize what to do (Participant 2)*

#### 3. Specific Barriers & Concerns

Participants were asked to consider barriers that they or colleagues might perceive around accessing an educational CPD intervention around burnout. Several participants highlighted perceived stigma associated with discussing mental health issues along with concerns that those in need may not have the insight to seek help, if in advanced stages of burnout:

> *there’s a bit of stigma and you don’t want to be seen, typically, as someone who rocks the boat, who moans, who seems to be critical about their work environment or the people they’re working with. Some people might prefer it to be anonymized. Some people don’t want to admit they’re struggling, maybe. Just reluctance to engage (Participant 5)*
>
> *I think there’ll be reluctance to admit to stress related conditions or burnout (Participant 4)*
>
> *you know when you think about the individual barriers that we might face, actually when you are, maybe, in that state of burnout and your outlook on things is going to be a little bit different (Participant 3)*
>
> *I think we’re all like “I need to do this, this and this” and then actually you don’t often have time to then start thinking about, “OK, I’ll spend some time to learn about methods, to reduce burnout”. And potentially by the time that you have burnt out, you’re kind of, and you realise you do need that, you’re probably it’s probably kind of too late to actually be doing that, isn’t it? (Participant 1)*

Participants also acknowledged that many GPs may be sceptical of a mindfulness-based or meditation-based intervention:

> *I think kind of you’re probably you pitching this at a group of people that may be sceptical, I suppose, and it I think explaining that there is evidence that supports its use would definitely be helpful for that, because they may then…it may be something that people are sceptical about, but just never really looked into um, because it’s not something that’s particularly interested them. And but yeah, so I think yeah, looking at kind of collating some evidence that kind of proves the use of it (Participant 1)*

Several participants also acknowledged that GPs may be concerned about admitting the true extent of their feelings due to the consequences for GMC referral or issues with their license to practise. This was felt to particularly be the case if the symptoms were being managed with alcohol or illicit drugs:

> *if you’re in severe burnout, in depression, there may be substances being used and, what have you, to manage these things. Particularly, you know, for a high functioning professional, like a GP, who is maybe worried about things like GMC and being reported to other organizations. And the knock-on impact it might have if there were questions about fitness to practice (Participant 3)*
>
> *You know, when you get GPs to come forth and participate, I think it’s a case of “Oh look out, you know what they’re looking for now” and the suspicion and paranoia and “who’s gonna know this and, you know, what are they going to do with it?”. And you know, I think there’ll be a barrier from that point of view, especially with the older GPs, I would imagine. So, I think that can be felt as a bit of a threat - to open up to such things. (Participant 5)*

Two participants raised potential issues of the term “resilience”, feeling that it has connotations suggesting the clinician is the problem for not being “resilient enough”, whilst ignoring the broken system within which the clinician is trying to work. It was felt that this may discourage GPs from engaging with a possible intervention:

> *I think the concept of “you need to become more resilient” can often feel quite difficult if it feels like you’re getting blamed for the problem, I think that might be a barrier (Participant 1)*
>
> *there’s also that question of like, well, you’re treating the problem, you’re treating the problem and not the cause or however that saying goes. So you know what everyone’s going to be is, like, “well, I’m burnt out because the system’s rubbish and, you know, I wouldn’t be if I could just do what I need for my patients”. But that’s the life, isn’t it? It’s not going to change anytime soon. So you might as well have some techniques to tackle it. (Participant 5)*

One of the participants noted that any intervention that would require time out of surgery would need to be adequately resourced, noting that simple monetary compensation, even paying for locum GP cover, could potentially be a barrier to uptake:

> *If you if you take the GP out the surgery and then you lose a session, you’re going to contribute to the burnout, you know, because your waiting list goes up and so you’ve got to be careful how, when there’s no resource. And it’s the lack of resource that leads to the burnout then you’re going to have to think carefully about it being adequately resourced. (Participant 4)*

## Discussion

### Main Findings

The objectives of the study were fully achieved. Two of five participants noted any prior exposure to CPD that focused on burnout or stress reduction. These were generic modules that were felt to be helpful for the purposes of introducing burnout as a concept. One participant noted two non-CPD based interventions put in place by previous colleagues including a positive feedback monthly newsletter and a daily lunchtime yoga/mediation session. Participants found the idea of a focused educational intervention to improve burnout to be a novel and beneficial idea that they would be happy to engage with. Participants noted the ideal format would be a face to face, group-based session broken up into smaller sessions over time. Some participants were opposed to an e-module format, but others saw the benefits of it in addition to a face-to-face intervention.

Participants highlighted several perceived barriers that they felt would need to be overcome in order for GPs to engage with a burnout intervention. Stigma of admitting feeling of burnout amongst colleagues was seen as a potential barrier and several participants raised concerns about GPs withholding the extent of their true feelings due to concerns over GMC referral. Scepticism over mindfulness and CBT based interventions was seen as a possible barrier, as well as the need for adequate resourcing to allow GPs to attend such an intervention. Several participants also highlighted frustration with suggesting GPs needed increased resilience, commenting that this could be seen as pointing the finger of blame on clinicians rather than identifying more significant system faults.

### Strengths & Limitations

To the best knowledge of the author, this is the first study of its kind to qualitatively explore attitudes of GPs towards CPD as it relates to burnout, assessing attitudes and perspectives on CPD interventions as a tool to help improve symptoms of burnout. All objectives of the study were achieved. A qualitative design allowed rich data to be captured, with an iterative interview approach allowing evolution of interview topics in a manner that a quantitative study design would not allow.

The study has several limitations. Due to time constraints, the assertion of data saturation could not be fully tested. Other themes may have emerged with further interviews of other participants. All of the participants were current or ex-colleagues of the author. This prior relationship may lead to bias, particularly regarding selection of participants. It may potentially influence participants’ responses to certain questions. All participants report working within NHS health boards in South Wales, therefore the results may not be generalisable to other locations.

All interviews and data analysis were completed by DW. Whilst this is likely to lead to greater uniformity of the data collection and analysis process, data analysis may have yielded different results if other researchers were involved in the data analysis process. The author is engaged in the annual appraisal process as a clinician, so his pre-existing views may bias interpretation of the participants data.

### Implications for Future Policy and Research

The results have provided rich and useful data to inform further research. The finding of the present study suggest that GPs would be keen to engage with educational CPD interventions to reduce burnout and improve wellbeing. An in-person face to face, small group format is likely to be most favoured design. A basic e-module introducing the concept of burnout with basic strategies to help, and signposting to other resources may also be of benefit. Several perceived barriers were noted which may prevent engagement with such an intervention and these would need to be considered in the design. Protected teaching afternoons, already funded by local GP clusters, may represent an ideal opportunity for these interventions. Further research would be needed to assess the effect of these interventions and whether they represent a cost-effective solution to the growing issue of burnout in the GP population.

Although not specifically enquired about during this study, several participants noted significant stresses and negative emotions associated with clinical work, in keeping with experiences of burnout. There has been no work to specifically quantify the prevalence of burnout in the Welsh GP population. This work would be useful to guide the need for further intervention and enable appropriate allocation funding in this area.

## Conclusion

GPs have a positive view of the potential for an educational CPD intervention to reduce burnout and improve wellbeing and have indicated a keenness to engage with such an intervention. Several important perceived barriers were highlighted that would need to be considered in the design and implementation of such an intervention. Further interventional research is required to assess the effectiveness of these interventions in a real-world scenario.

## Data Availability

All data produced in the present study are available upon reasonable request to the authors

## Notes

### Competing Interest Statement

The authors have declared no competing interest.

### Funding Statement

This study did not receive any funding

### Author Declarations

Swansea University Medicine, Health and Life Science Ethics Approval Board of Swansea University gave ethical approval for this work

